# Derivation and Validation of Clinical Prediction Rule for COVID-19 Mortality in Ontario, Canada

**DOI:** 10.1101/2020.06.21.20136929

**Authors:** David N. Fisman, Amy L. Greer, Ashleigh R. Tuite

## Abstract

**Background:** SARS-CoV-2 is currently causing a high mortality global pandemic. However, the clinical spectrum of disease caused by this virus is broad, ranging from asymptomatic infection to cytokine storm with organ failure and death. Risk stratification of individuals with COVID-19 would be desirable for management, prioritization for trial enrollment, and risk stratification. We sought to develop a prediction rule for mortality due to COVID-19 in individuals with diagnosed infection in Ontario, Canada.

**Methods:** Data from Ontario’s provincial iPHIS system were extracted for the period from January 23 to May 15, 2020. Both logistic regression-based prediction rules, and a rule derived using a Cox proportional hazards model, were developed in half the study and validated in remaining patients. Sensitivity analyses were performed with varying approaches to missing data.

**Results:** 21,922 COVID-19 cases were reported. Individuals assigned to the derivation and validation sets were broadly similar. Age and comorbidities (notably diabetes, renal disease and immune compromise) were strong predictors of mortality. Four point-based prediction rules were derived (base case, smoking excluded as a predictor, long-term care excluded as a predictor, and Cox model based). All rules displayed excellent discrimination (AUC for all rules <u>> 0.92</u>) and calibration (both by graphical inspection and P > 0.50 by Hosmer-Lemeshow test) in the derivation set. All rules performed well in the validation set and were robust to random replacement of missing variables, and to the assumption that missing variables indicated absence of the comorbidity or characteristic in question.

**Conclusions:** We were able to use a public health case-management data system to derive and internally validate four accurate, well-calibrated and robust clinical prediction rules for COVID-19 mortality in Ontario, Canada. While these rules need external validation, they may be a useful tool for clinical management, risk stratification, and clinical trials.

## Introduction

Since the COVID-19 pandemic was declared by the World Health Organization on March 12, 2020 (1), the spread of SARS-CoV-2 has taken a fearsome toll on global mortality. As of June 11, 2020, over 400,000 deaths worldwide have been attributed to SARS-CoV-2, with many more excess deaths likely related either to infection with the virus or disruption of health systems by epidemics (2). While most infections with SARS-CoV-2 are mild or even asymptomatic, approximately 20% of recognized infections are sufficiently severe to require hospitalization (3, 4). Among those hospitalized, 10-20% have an intensive care requirement, usually related to respiratory failure (3-5), though multiorgan system failure (6), clotting abnormalities (7) and angioneogenesis (8) with resultant bleeding are increasingly recognized as severe complications of COVID-19.

Numerous studies have identified clinical factors associated with requirements for intensive care and death among those with COVID-19 infection (9-11). Published prediction models to date have evaluated case-level factors that might predict care diagnosis, more severe disease requiring hospitalization, and poor outcomes (critical illness or death) (9). A recent review identified 16 prediction models focused on prognosis; 14 were based on the COVID-19 epidemic in China and the other two used aggregated public data from a variety of sources (9). The generalizability of these rules to the North American context is unclear. Furthermore, few of these efforts included conversion of prediction models into parsimonious, simple, score-based tools that can be used easily for risk stratification in clinical settings. In the context of COVID-19, a such rules might have important implications for risk-stratification of patients, streamlining decisions around hospital care vs. self-isolation (12), and prioritizing individuals for enrollment in clinical trials of emerging therapies (e.g., convalescent plasma or antiviral drugs), as has been the case with similar tools developed for community acquired pneumonia (13).

Ontario, Canada, had identified over 30,000 virologically confirmed cases of COVID-19 in the province as of June 11, 2020 (14). Each confirmed case is the subject of epidemiological investigation by local public health authorities, who enter epidemiological, clinical and outcome data into the Province’s Integrated Public Health Information System (iPHIS). Our objective was to make use of iPHIS data to develop and validate parsimonious, sensitive and specific prediction rules for infection-related death in individuals with COVID-19 in Ontario.

## Methods

### Study population and data collection

Ontario is Canada’s most populous province, with a current population of 14.7 million (15). The Province identified imported COVID-19 cases from China, and Iran, in January and February 2020 (16); local epidemic spread of SARS-CoV-2 has been evident since late February 2020 (17). Each of Ontario’s 34 public health units is responsible for local case investigation and uploading of case information into the iPHIS data system, which is used for surveillance and case management of notifiable diseases in the Province (18). Ontario’s case definition for a confirmed case of requires a positive laboratory test using a validated nucleic acid amplification test, including real-time PCR and nucleic acid sequencing (19).

Information on patient characteristics – including age group (by 10-year intervals), sex, medical comorbidities, long-term care residence, healthcare and emergency service work, case symptoms, dates of symptom onset, testing and reporting, hospitalization and intensive care admission, and mortality was collected for cases. Approximately 80% of all deaths during the Ontario COVID-19 epidemic have occurred in long term care facilities (20), and there has been little transfer of long-term care residents to intensive care units (17).

### Statistical analysis

We randomly assorted cases into derivation and validation sets. Characteristics of the two sets are presented in **Table 1**. Univariable logistic regression was used to identify factors associated with mortality in the derivation group. Continuous variables were dichotomized to facilitate score generation and ease of application in clinical settings. When a factor was found to be protective, the covariate evaluated was *absence* of the factor, so that resultant odds ratios were > 1.

**Table 1.** Characteristics of Confirmed COVID-19 Cases in Ontario, Canada to May 15, 2020.

| Covariate | Overall (% or Median, IQR) | Derivation (% or Median, IQR) | Validation (% or Median, IQR) | P-value |
| --- | --- | --- | --- | --- |
| All | 21922 (100) | 10957 (50) | 10965 (50) |  |
| Age > 59 | 9398 (43) | 4672 (43) | 4705 (43) | 0.63 |
| FSA Income* | \$64,869 (\$26,402) | \$64,986 (\$26,937) | \$64,869 (\$26,402) | 0.73 |
| Male gender | 9389 (43) | 4708 (43) | 4681 (43) | 0.68 |
| Time from symptom onset to report | 5 (6) | 5 (6) | 5 (6) | 0.81 |
| Long-term care resident | 3102 (14) | 1539 (14) | 1563 (14) | 0.50 |
| Outbreak-associated case<sup>†</sup> | 9438 (43) | 4772 (44) | 4666 (43) | 0.14 |
| Healthcare worker | 3780 (17) | 1888 (17) | 1892 (17) | 0.55 |
| Homeless shelter worker | 106 (0.4) | 61 (0.6) | 45 (0.4) | 0.13 |
| Homeless | 226 (1) | 102 (0.9) | 124 (1) | 0.11 |
| Smoker (recorded) | 515 (2) | 285 (3) | 230 (2) | 0.01 |
| Pregnant or post-partum | 91 (0.4) | 58 (0.5) | 45 (0.4) | 0.21 |
| Comorbidity history |  |  |  |  |
| Anemia or hemoglobinopathy | 370 (2) | 177 (2) | 193 (2) | 0.42 |
| Chronic liver disease | 94 (0.4) | 38 (0.3) | 56 (0.5) | 0.06 |
| Renal disease | 358 (2) | 185 (2) | 173 (2) | 0.55 |
| Diabetes | 1294 (6) | 623 (6) | 671 (6) | 0.22 |
| Chronic obstructive pulmonary |  |  |  |  |
| disease | 267 (1) | 134 (1) | 133 (1) | 0.97 |
| Asthma | 880 (4) | 409 (4) | 390 (4) | 0.58 |
| Cardiovascular disease | 2032 (9) | 979 (9) | 1053 (10) | 0.15 |
| Malignancy | 460 (2) | 211 (2) | 249 (2) | 0.06 |
| Immune compromise | 318 (1) | 162 (1) | 156 (1) | 0.69 |
| Tuberculosis | 52 (0.2) | 32 (0.2) | 20 (0.2) | 0.10 |
| Obesity | 295 (1) | 137 (1) | 158 (1) | 0.16 |
| Outcomes |  |  |  |  |
| Hospitalized | 2779 (13) | 1355 (12) | 1424 (13) | 0.17 |
| Record of intubation and/or |  |  |  |  |
| mechanical ventilation | 408 (2) | 195 (2) | 213 (2) | 0.31 |
| Died | 1825 (8) | 862 (8) | 963 (9) | 0.02 |

**NOTE:** Proportions compared with chi-squared test, continuous variables compared with Wilcoxon rank-sum test.
\*Based on mean after tax income by FSA (2016 Canadian Census).
†Defined as case or cases with outbreak number signifying part of an outbreak investigation by a public health unit.

Risk factors significant at P < 0.2, or which were thought a priori to confer important increases in risk (age and sex) were included in model building using a forward stepwise selection algorithm, with covariates selected for P < 0.05, and retained in the model for P < 0.15. We did not include interaction terms in efforts to keep a final prediction rule as simple as possible. The final regression model was transformed to a point-based rule, with each regression coefficient divided by half of the smallest coefficient and rounded to the nearest integer to obtain weighted values. Risk scores were calculated by summing the individual point values of all applicable risk factors. Risk of death can then be approximated from a graph of model-predicted probability versus calculated score (**Figure 1**) using the relation p = 1/(e^-(I + CS)^+1) where S is the individual’s score, C is the prediction rule’s coefficient in a logit model using score as a predictor of death, I is the intercept from the same model.

**Figure 1.**
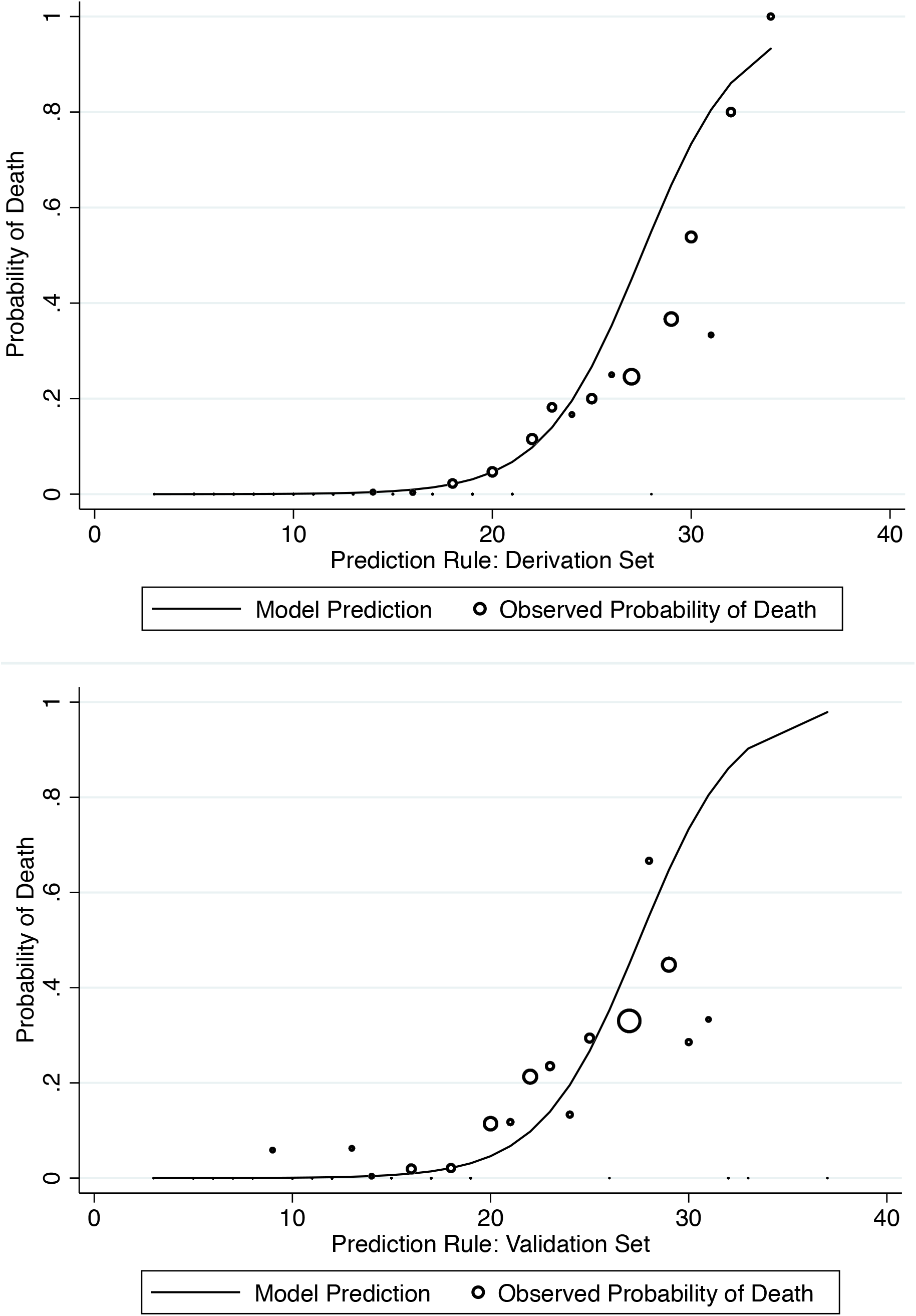
Observed and Predicted Risk of Death by Score, Base-Case Rule. Plot of predicted probability of death (Y-axis) by model score (X-axis) for base case prediction rule. Curve represents model predictions, circles represent observed proportion who died. Circle size proportionate to number of deaths at a given score. Top panel: derivation set; bottom panel: validation set.

The discriminatory ability of the prediction rule in the derivation group was quantified through the area under the receiver-operating characteristic curve (ROC AUC), with 95% confidence intervals estimated through 1000 bootstrap replicates. Calibration was assessed visually and using the Hosmer-Lemeshow test for goodness of fit, which evaluates expected and observed probabilities in population deciles (21).

### Survival Analytic Approach and Alternate Rules

Some analysts have expressed concern that failure to account for right censoring in could lead to bias in COVID-19 clinical prediction rules (9). As such we created a second prediction rule using Cox-proportional hazards analysis, by identifying factors associated with increased hazard of death using the same selection algorithm as applied to the logistic model described above. Log transformed hazard ratios were converted to point scores using the approach described above. Discriminative ability of the rule was evaluated using Harrell’s C-statistic after constructing a Cox proportional hazards model with the score as the sole covariate in both the derivation and validation sets. ROC analysis, and score calibration, were performed by using the Cox-model-derived score as a predictor in a logistic model.

Smoking status emerged as a protective effect in our base case prediction model; this is likely to be controversial with some users. Furthermore, it might be argued that the known high mortality associated with COVID-19 in long term care settings favors creation of a rule for non-long-term care residents. As such, we made additional rules which excluded smoking status, and which excluded long-term care residents, using the approach described above.

### Sensitivity Analyses

In the base case, models were built using only observations from individuals with complete data; we tested the robustness of our models by evaluating the discriminative ability and calibration of rules in datasets in which missing fields were replaced at random, and in datasets where an attribute was assumed not present if a field was left blank (e.g., if an individual had no record of presence or absence cardiac disease, they were assumed not to have cardiac disease). All analyses were performed using Stata version 14.0 (Stata Corporation, College Station, TX). The study was approved by the research ethics board of the University of Toronto.

## Results

Of 21,922 COVID-19 cases reported between January 23 and May 15, 2020, 57% were female, and 43% were aged > 59 years. The median time from symptom onset to case reporting was 5 days (IQR 4 to 10 days). Fourteen percent of cases were residents of long-term care facilities; 17% were healthcare workers. Thirteen percent of cases were hospitalized; 2% had record of intubation and/or mechanical ventilation, and case-fatality was 8%. Individuals assigned to the derivation and validation sets were broadly similar, but were significantly more likely to be smokers, less likely to have a history of chronic liver disease, and less likely to die (**Table 1**).

### Derivation of the Prediction Rule

In univariable analyses, death was associated with a broad array of demographic characteristics and comorbid conditions. No association was seen between risk of death and mean neighborhood income or asthma which were not included in subsequent model building (**Table 2**). As age was provided as ordinal, 10-year age groupings (0 to 9, 10 to 19, 20 to 29, etc.) the age coefficient in models represents increased risk per increase in (age/10). Using a forward selection algorithm, we identified 7 independent predictors of death in the derivation group: age, long-term care residence, a history of renal disease, diabetes, chronic obstructive pulmonary disease, and immune compromise, and non-smoking. (**Table 3**).

**Table 2.** Univariable and Multivariable Analyses, and Point Score Derivation, Base Case Prediction Rule.

| Covariate | Univariable OR (95% CI) | P-value | Multivariable OR (95% CI) | Logit | Points |
| --- | --- | --- | --- | --- | --- |
| Age (per 10-year increment) | 3.48 (3.28 to 3.70) | <0.001 | 2.42 (1.78 to 3.29) | 0.88 | 2 |
| Low income* | 1.04 (0.92 to 1.18) | 0.55 | --- | --- | --- |
| Male gender | 1.13 (1.02 to 1.25) | 0.02 | --- | --- | --- |
| Time from symptoms to diagnosis $\leq 3$ days<sup>†</sup> | 1.27 (1.14 to 1.42) | <0.001 | --- | --- | --- |
| Long-term care resident | 22.62 (19.08 to 26.83) | <0.001 | 6.24 (2.95 to 13.21) | 1.83 | 4 |
| Outbreak-associated case | 9.15 (8.10 to 10.33) | <0.001 | --- | --- | --- |
| Non-healthcare worker<sup>‡</sup> | 30.56 (15.77 to 59.22) | <0.001 | --- | --- | --- |
| Non-homeless shelter worker<sup>‡</sup> | 5.79 (0.80 to 41.96) | 0.08 | --- | --- | --- |
| Non-homeless<sup>‡</sup> | 2.31 (0.94 to .712143) | 0.07 | --- | --- | --- |
| Non-smoker<sup>‡</sup> | 1.65 (0.98 to 2.77) | 0.06 | 6.86 (0.73 to 64.27) | 1.93 | 4 |
| Pregnant or post-partum | No deaths | --- | --- | --- | --- |
| Comorbidity history |  |  |  |  |  |
| Anemia or hemoglobinopathy | 5.08 (3.68 to 7.02) | <0.001 | --- | --- | --- |
| Chronic liver disease | 6.06 (3.50 to 10.46) | <0.001 | --- | --- | --- |
| Renal disease | 9.85 (7.31 to 13.26) | <0.001 | 2.37 (0.97 to 5.77) | 0.86 | 2 |
| Diabetes | 6.49 (5.22 to 8.06) | <0.001 | 2.19 (1.08 to 4.42) | 0.78 | 2 |
| Chronic obstructive pulmonary disease | 11.22 (8.14 to 15.44) | <0.001 | 3.26 (1.15 to 9.26) | 1.18 | 3 |
| Asthma | 1.01 (0.71 to 1.44) | 0.96 | --- | --- | --- |
| Cardiovascular disease | 11.38 (9.12 to 14.20) | <0.001 | --- | --- | --- |
| Malignancy | 6.36 (4.80 to 8.44) | <0.001 | --- | --- | --- |
| Immune compromise | 4.12 (2.94 to 5.79) | <0.001 | 3.56 (1.12 to 11.35) | 1.27 | 3 |
| Tuberculosis | 0.88 (0.21 to 3.70) | <0.001 | --- | --- | --- |
| Obesity | 2.63 (1.78 to 3.89) | <0.001 | --- | --- | --- |

\*Residence in FSA in lowest quartile of income
†Lowest quartile lag between symptoms and diagnosis
‡Non-exposure status evaluated as risk factor to maintain positive covariate.

**Table 3.** Base Case and Alternate Clinical Prediction Rules.

| Covariate | Rule 1:<br>Base Case | Rule 2: Cox<br>model-based* | Rule 3: Non-<br>smokers Excluded | Rule 4: Long term care<br>residents excluded |
| --- | --- | --- | --- | --- |
| Age | 2 | 3 | 3 | 2 |
| Male sex | --- | 2 | --- | --- |
| Renal disease | 2 | --- | 2 | 3 |
| Immune compromised | 3 | 4 | 5 | 4 |
| Diabetic | 2 | 4 | 3 | 2 |
| COPD | 3 | 3 | 3 | --- |
| Cardiovascular disease | --- | --- | 2 | 4 |
| Long-term care resident | 5 | --- | 7 | --- |
| Non-smoker | 5 | --- | --- | --- |
| Time from symptoms to<br>diagnosis $\leq 3$ days | --- | --- | --- | 2 |
| Maximum points | 40 | 40 | 50 | 40 |
| Smallest logit model coefficient<br>(C)* | 0.36 | 0.34 | 0.25 | 0.52 |
| Model intercept (I)* | -9.81 | -9.99 | -8.33 | -12.51 |
| AUC in derivation set (validation set) | 0.95 (0.92) | 0.93 (0.91) | 0.95 (0.92) | 0.92 (0.91) |
| Hosmer-Lemeshow goodness-of-fit test P-value in derivation set (validation set) | 0.85 (0.20) | 0.50 (0.24) | 0.99 (0.40) | 0.59 (0.04) |

**NOTE:** AUC, area under the ROC curve; Hosmer-Lemeshow test based on deciles of risk score.
\* Can be used to calculate probability of death as per text.

The point-based prediction rule was well-calibrated between quantiles of observed and expected risk (Hosmer-Lemeshow χ^2^=1.58; p=0. 0.90) in the derivation group and discriminated extremely well between those who did and did not develop die (ROC AUC in the derivation group=0.95; 95% CI, 0.91-0.96). The median score (interquartile range) was 13 (6) for survivors and 25 (6) for those who died (p<0.001 by the Wilcoxon rank-sum test). The rule displayed good calibration to outcomes in the validation set (Hosmer-Lemeshow χ^2^=9.16; p=0.16), as well as excellent discrimination (AUC 0.92, 95% CI 0.89 to 0.94) (**Figure 1** and **Figure 2**).

**Figure 2.**
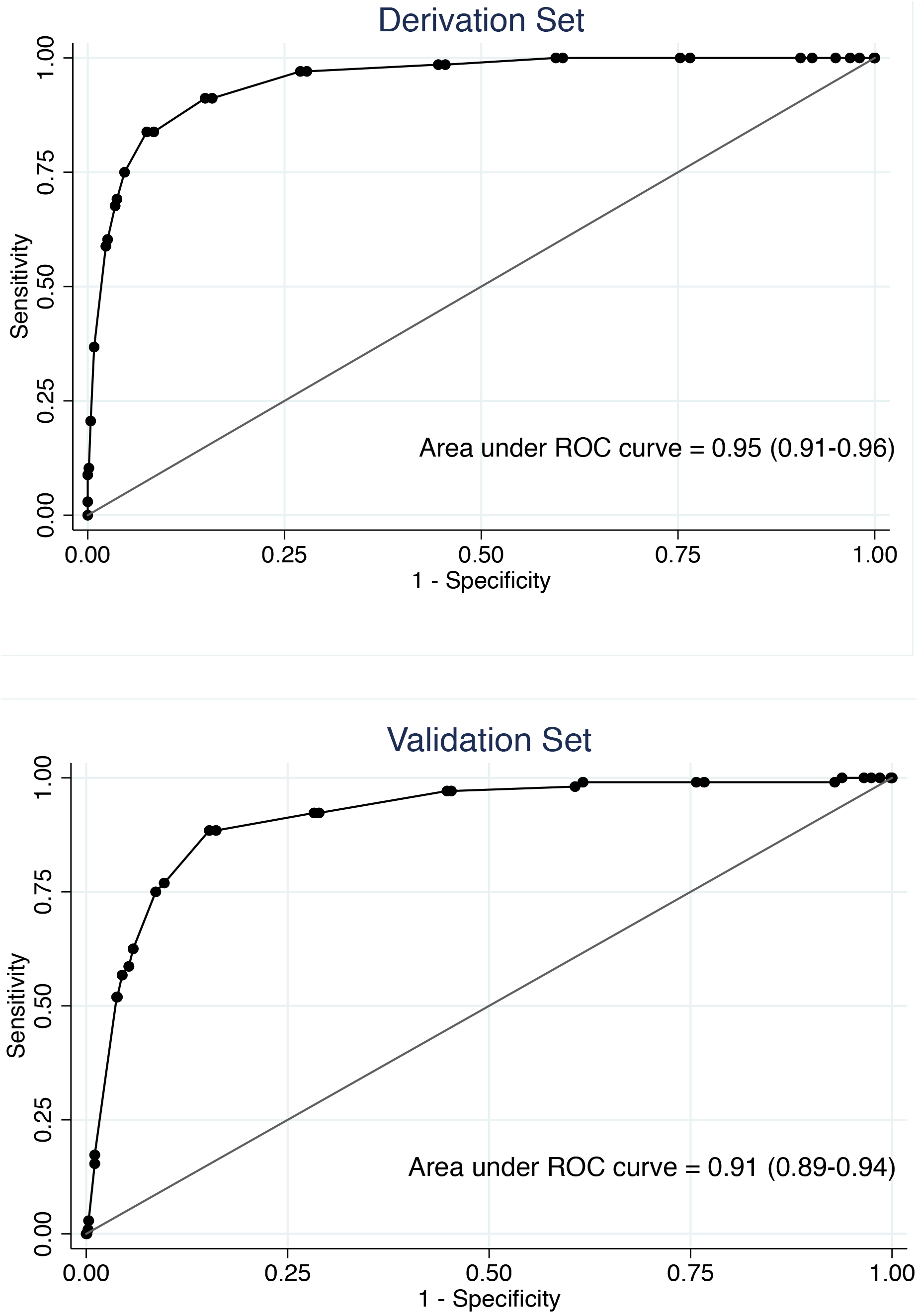
Receiver Operator Characteristic Curve, Base-Case Rule. Sensitivity of rule (Y-axis) is plotted against false positive rate (1-specificify, X-axis) for different positivity criteria available from score. Confidence intervals for area under the curve derived via boostrapping. Top panel: derivation set; bottom panel: validation set.

### Alternate Prediction Rules

Three alternate rules (based on a Cox proportional hazards model, a logistic model excluding smoking status, and a model with long-term care residents excluded) were created. These models had excellent discrimination. We statistical found evidence for poor calibration of the model that excluded long-term care residents in the validation set (P = 0.04). The Harrell’s C-statistic for a Cox model including age, male sex, diabetes, COPD, and immune compromise was 0.97 in the derivation set, and 0.96 in the validation set. Other fit statistics, and c-statistics for AUC, as well as values of the model intercept and smallest logit model coefficient (for calculation of death probability) are presented in **Table 3** and presented graphically in the **Supplement**.

### Sensitivity Analyses

We re-evaluated all four prediction rules in datasets in which missing variables were assumed to not be present, and in which missing variables were replaced randomly. Discriminative ability remained good for both randomly replaced datasets (ROC curve AUC 0.84-0.90 for missing observations replaced with zeroes; AUC 0.79-0.83 for missing observations replaced randomly). The large number of observations in datasets with all missings replaced (N = 21,922) resulted in statistically significant differences between observed and expected mortality probabilities (P < 0.001 for all analyses by Hosmer-Lemeshow test), but visual inspection suggested that calibration of rules remained very good (**Supplement**).

## Discussion

Accurate prediction of mortality from COVID-19 has a number of potential applications, including rational decision making for hospital admission, prioritization of high-risk individuals for inclusion in trials of novel therapeutic agents, and to identify high risk individuals for policy purposes (e.g., to inform decisions around risks and benefits of remote work). We demonstrate here that COVID-19 mortality in identified cases can be predicted with remarkable accuracy based on the limited, readily available demographic and chronic health information available in public health line lists. The large number of COVID-19 cases that have occurred in Ontario provided sufficient statistical power for both model derivation and validation without resorting to bootstrap resampling. The discriminative ability of our rules (as reflected in AUC > 0.9 in both derivation and validation sets) places them among the upper tier of current COVID-19 prediction rules; the parsimoniousness of these rules and their conversion to an easy-to-calculate point score allows easy incorporation into clinical care.

While many of our predictors (age and comorbidities) could have been anticipated based on established epidemiology of COVID-19 (22-24), some (e.g., non-smoking as a predictor of mortality) are likely to be controversial, and it is for this reason that we derived alternate rules that exclude non-smoking. Apparent protective effects of smoking against COVID-19 acquisition (25) as well as under-representation of smokers among COVID-19 patients have been noted by others (26). However, other investigators have suggested higher risk of progression of COVID-19 in smokers(26, 27), and increased density of ACE-2 (a viral receptor) is present in the lungs of smokers (28), suggesting that apparent protective effects might result from selection bias (e.g., individuals predisposed to very mild COVID-19 infection as a result of young age or good general health might be over-represented among those tested for COVID-19 due to smoking-related health concerns like cough). Regardless, a non-causal association with risk may still be useful for clinical prediction; if this association reflects peculiarities of Ontario’s approach to COVID-19 testing we expect that it may not be generalizable to other jurisdictions that test more widely.

Similarly, the strong effect of long-term care residence on mortality is unsurprising, given the high fraction of long-term care deaths seen during the Canadian COVID-19 epidemic to date (20). As such we created alternate rules that exclude smoking and long-term care residence; these rules can be used in place of our base case rule, as they have similar discriminative ability. Lastly, to avoid biases that might be introduced by right-censoring (i.e., lack of mortality in individuals in the study cohort as a result of insufficient follow up time) we derived an additional rule using survival methods, which also performed well. There was substantial overlap between all four prediction rules in included covariates: notably, age, diabetes, and immune compromise were included in all four rules we derived, and renal or chronic obstructive pulmonary disease were included in 3 of four rules.

Our analysis had many limitations; the use of a public health record system not explicitly designed as a research tool means that we lack laboratory and radiological results that have been useful in other prediction models (23, 29). Furthermore, missing data was a significant limitation of our dataset, although our models appeared robust even with random replacement of predictors and outcomes that should bias associations towards the null. In that sense, the ability to derive simple, accurate and parsimonious rules, which perform well in split-halves validation, despite limitations in our dataset, may suggest generalizability of application outside Ontario. We hope that other groups will evaluate our rules in other settings.

In summary, we developed and internally validated a prediction rule for COVID-19 mortality using a large and detailed public health line list in the Canadian province of Ontario. The rule was well calibrated and discriminated well and was robust in sensitivity analyses to assess the impact of missing information on predictor variables. If externally validated, this rule might facilitate decision making during future epidemic waves.

## Data Availability

Primary data are unfortunately not available by agreement with the Government of the Province of Ontario.

## Supplement: Derivation and Validation of Clinical Prediction Rule for COVID-19 Mortality in Ontario, Canada

### 1. Cox-model derived prediction rule

As noted in the text, we used a Cox proportional hazards model to derive point scores for a prediction score. The use of Cox models was intended to avoid bias that might be introduced as a result of incomplete follow up and right censoring. Point scores were derived based on log-hazard ratios. The scores, and values for I and C necessary to predict mortality probability are presented in Table 3. The score itself was then evaluated as a single covariate in a logit model predicting mortality. Model calibration in derivation (top panel) and validation (bottom panel) sets are presented in Figure S1 below. Figure S2 presents the ROC curves for the derivation (top panel) and validation (bottom panel) sets.

**Figure S1.**
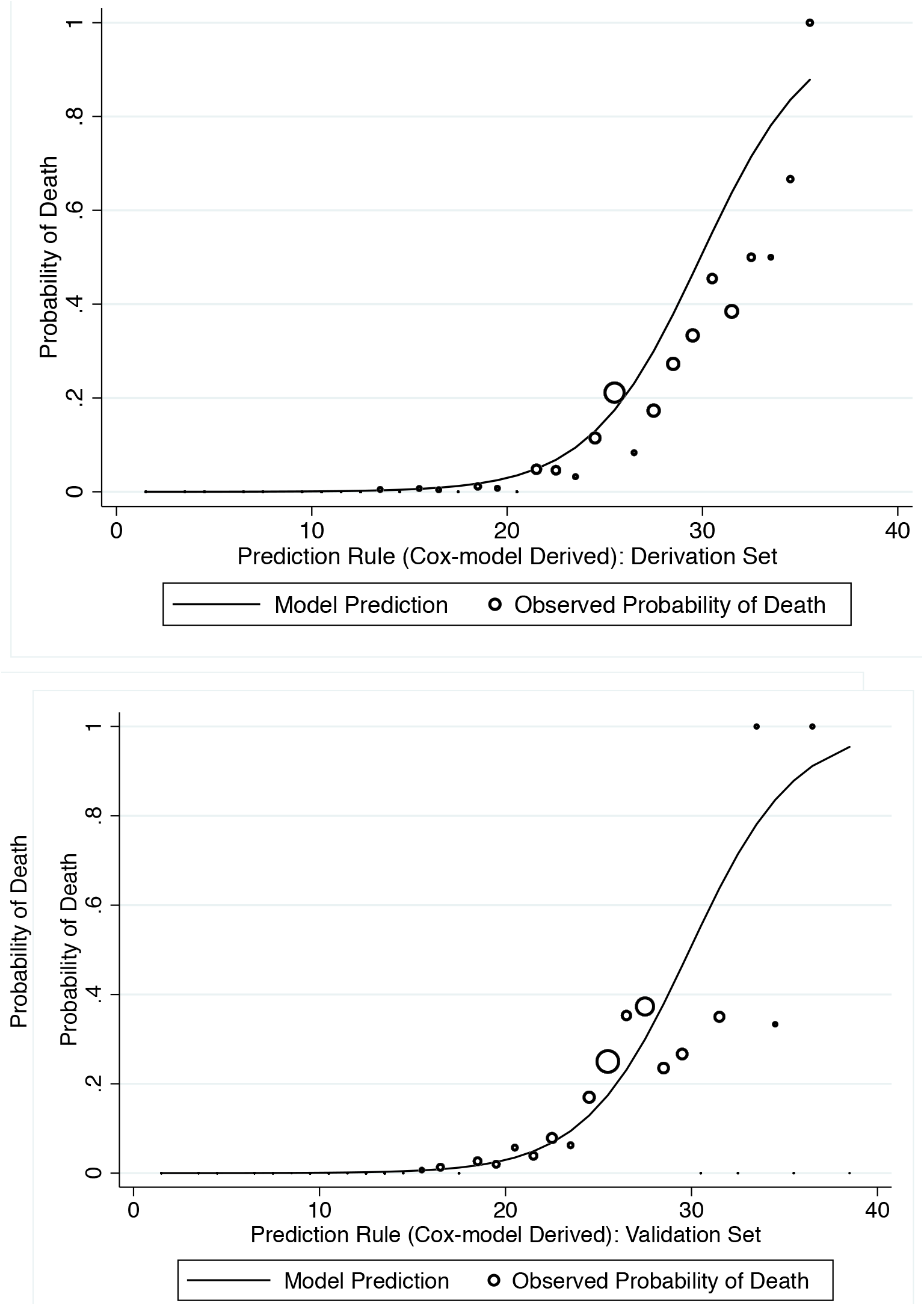
Observed (circles) and predicted (curve) probability of death in Cox model-derived alternate prediction rule. Circle size is proportionate to number of deaths for each score.

**Figure S2.**
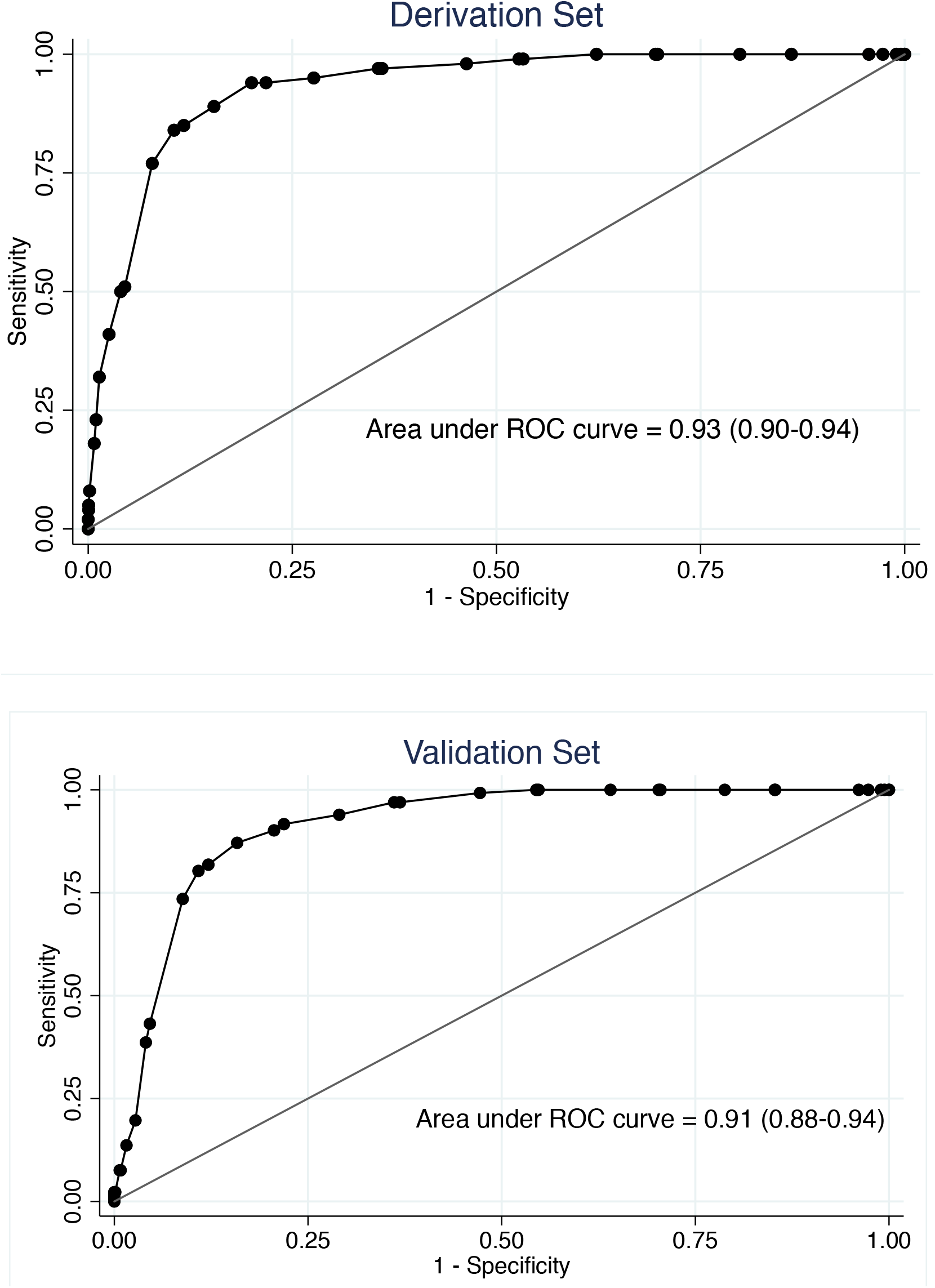
ROC curves for derivation (top) and validation (bottom) sets, Cox model-based prediction rule. Confidence intervals derived via bootstrapping.

### 2. Logit model-derived prediction rule with smoking excluded

We anticipated that a prediction rule incorporating non-smoking as a risk factor for mortality would be controversial. As such, we created an alternate rule with non-smoking excluded. Again, rule-based scores, and values for I and C necessary to predict mortality probability are presented in Table 3. As above model calibration in derivation (top panel) and validation (bottom panel) sets are presented in Figure S3 below. Figure S4 presents the ROC curves for the derivation (top panel) and validation (bottom panel) sets.

**Figure S3.**
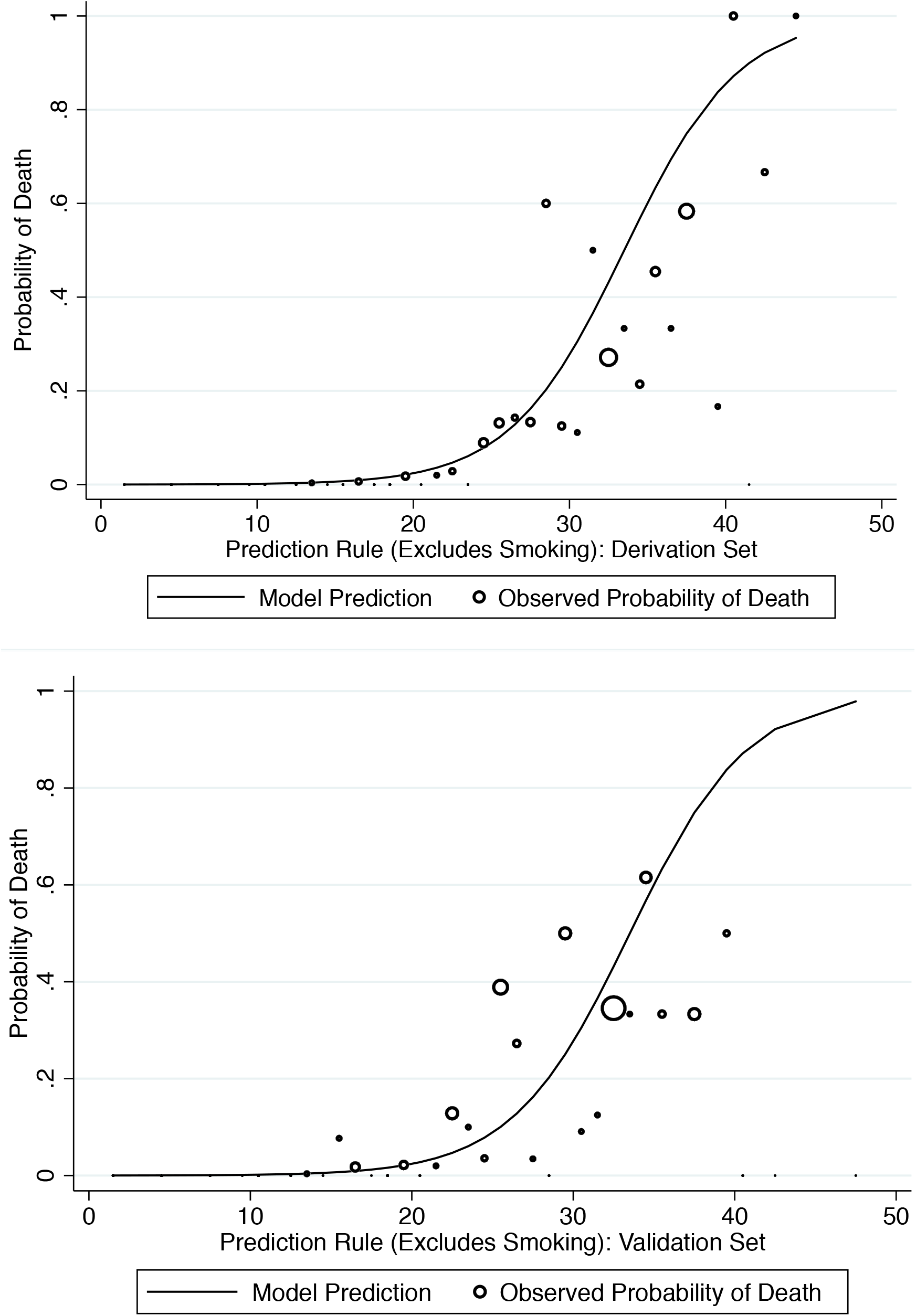
Observed (circles) and predicted (curve) probability of death in logit model-derived alternate prediction rule that excludes smoking status. Circle size is proportionate to number of deaths for each score.

**Figure S4.**
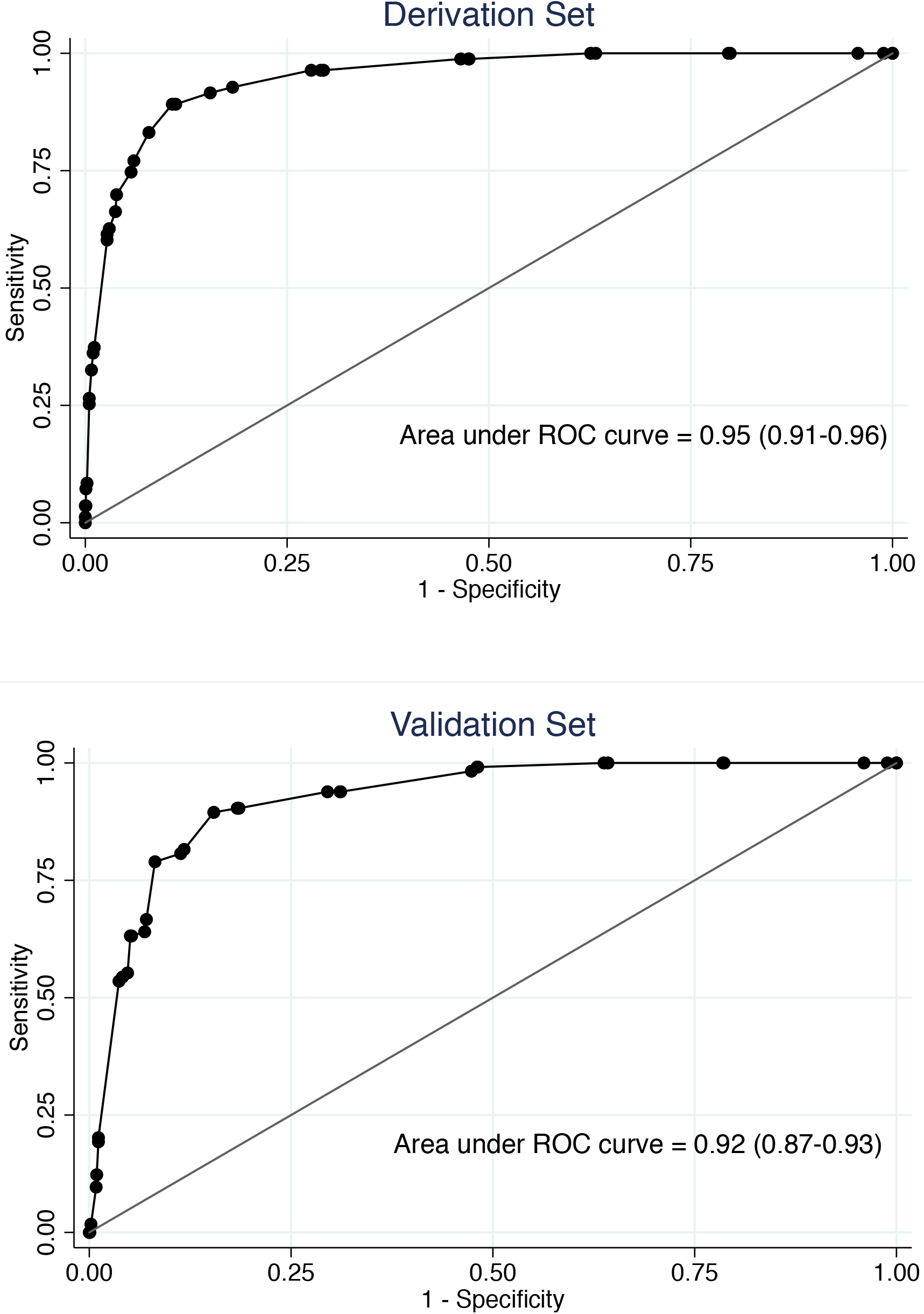
ROC curves for derivation (top) and validation (bottom) sets in logit model-derived alternate prediction rule that excludes smoking status. Confidence intervals derived via bootstrapping.

### 3. Logit model-derived prediction rule with long-term care residents excluded

The extremely high mortality in the long-term care setting might make a prediction rule unhelpful. As such, we created an alternate rule with long-term care residents excluded. Again, rule-based scores, and values for I and C necessary to predict mortality probability are presented in Table 3. As above model calibration in derivation (top panel) and validation (bottom panel) sets are presented in Figure S5 below. Figure S6 presents the ROC curves for the derivation (top panel) and validation (bottom panel) sets.

**Figure S5.**
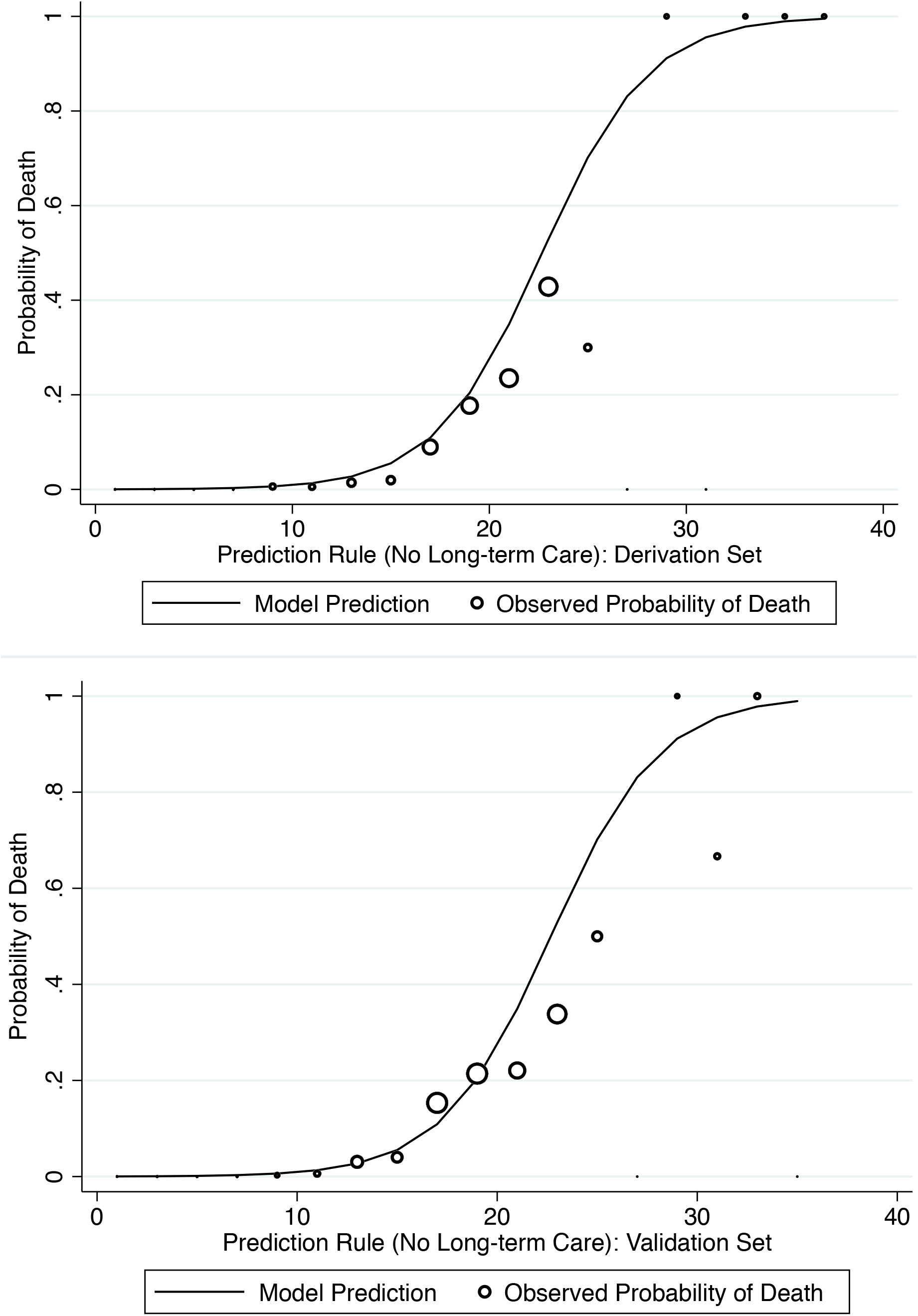
Observed (circles) and predicted (curve) probability of death in logit model-derived alternate prediction rule that excludes long term care residents. Circle size is proportionate to number of deaths for each score.

**Figure S6.**
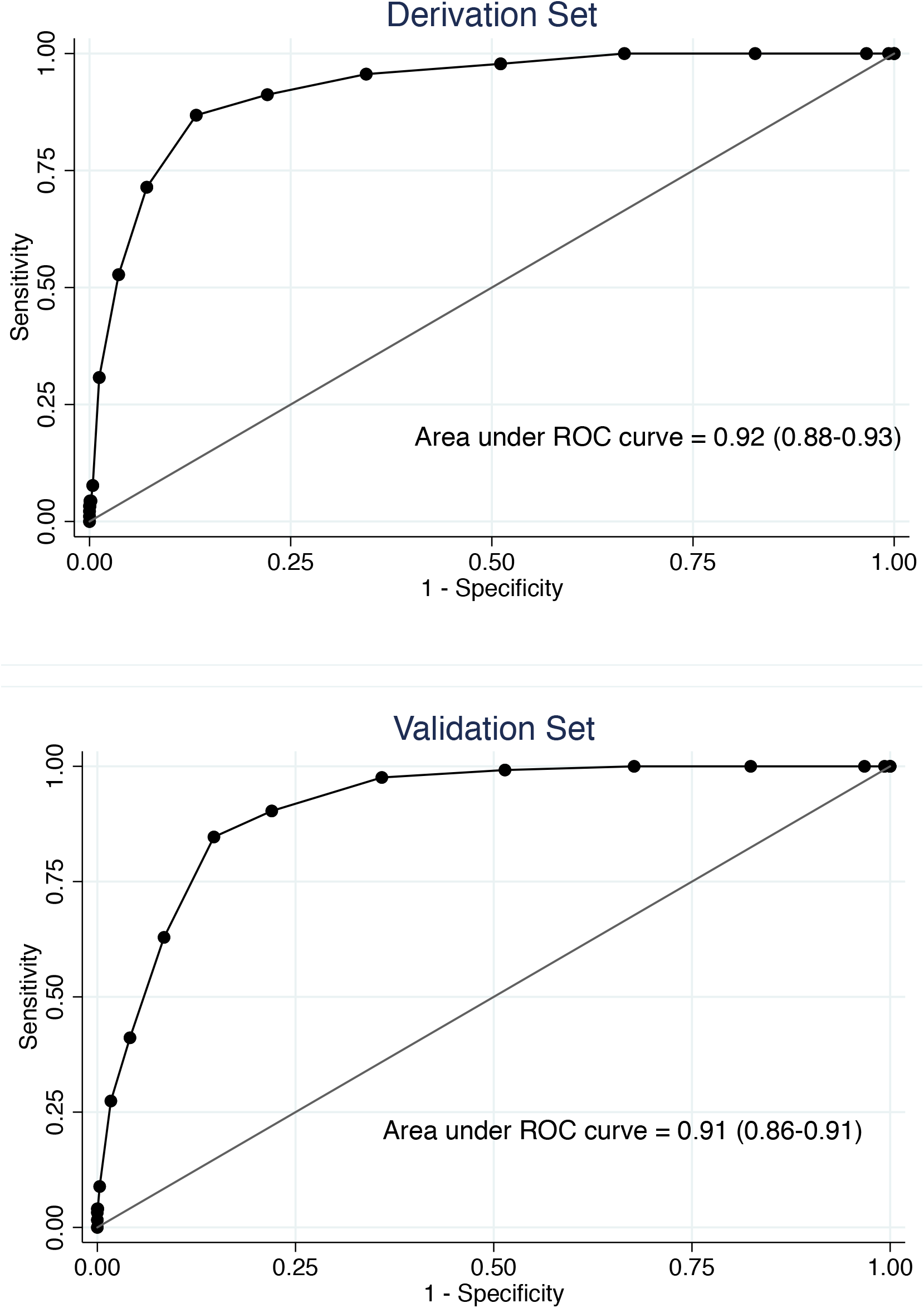
ROC curves for derivation (top) and validation (bottom) sets in logit model-derived alternate prediction rule that excludes smoking status. Confidence intervals derived via bootstrapping.

### 4. Sensitivity Analyses with Replacement of Missing Data

The iPHIS dataset was limited by substantial data missingness. To assess the robustness of our rules, we replaced missing variables in two ways: first, by assuming that missingness (for covariates or death) indicated that they were not present or did not occur (i.e., replaced missing values as zero); and second, by assuming that variables were missing completely at random, and replacing missing values randomly based on their frequency of observation among non-missings. The latter approach had the effect of substantially increasing the number of deaths available in the dataset. Notwithstanding the likely introduction of misclassification via both of these approaches, model calibration (based on visual inspection) and discrimination remained very good. Calibration (left sided panels) and discrimination (right sided panels) are presented graphically for all four prediction rules with missing values replaced as zeroes (Figure S7) and missing values replaced at random (Figure S8).

**Figure 7S.**
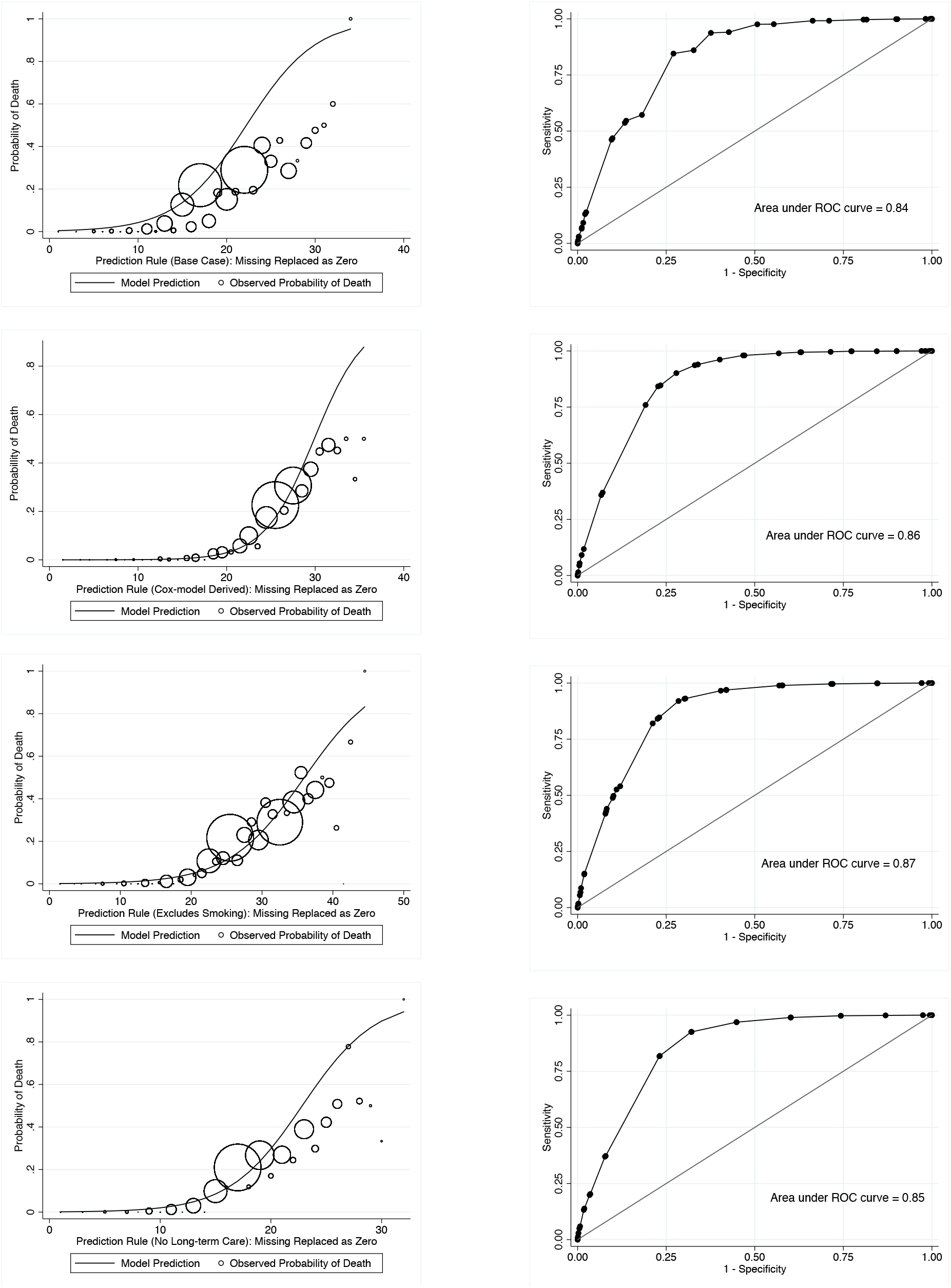
Left panel figures present model calibration for all four prediction rules with missing variables replaced as zeroes. Lines represent model predictions and circles represent observed probability of death; circle size is proportionate to observed numbers of deaths at each score level. Right panel figures represent ROC curves for the same prediction rules with missing variables replaced as zeroes.

**Figure 8S.**
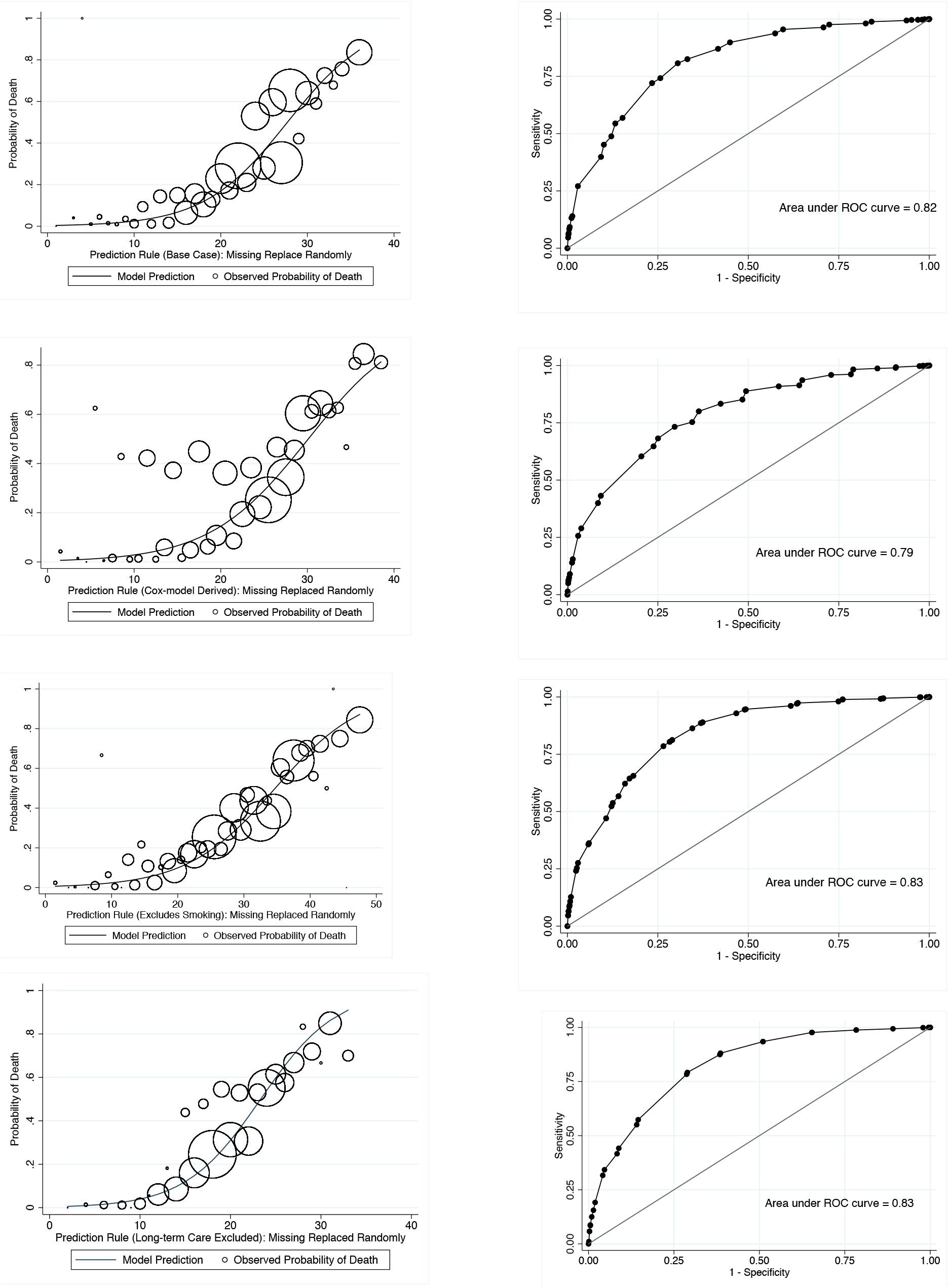
Left panel figures present model calibration for all four prediction rules with missing variables replaced at random. Lines represent model predictions and circles represent observed probability of death; circle size is proportionate to observed numbers of deaths at each score level. Circles are large because missing death data has been replaced as either present or absent, with the result that death numbers are far higher than in other analyses. Right panel figures represent ROC curves for the same prediction rules with missing variables replaced at random.

## References

1. World Health Organization. WHO announces COVID-19 outbreak a pandemic. Available via the Internet at euro.who.int/en/health-topics/health-emergencies/coronavirus-covid-19/news/news/2020/3/who-announces-covid-19-outbreak-a-pandemic. Last accessed June 14, 2020. 2020.

2. Johns Hopkins University. Coronavirus resource tracker. Available via the Internet at https://coronavirus.jhu.edu/data/new-cases. Last accessed June 20, 2020. 2020.

3. Wang D, Hu B, Hu C, Zhu F, Liu X, Zhang J, et al. Clinical Characteristics of 138 Hospitalized Patients With 2019 Novel Coronavirus-Infected Pneumonia in Wuhan, China. JAMA. 2020.

4. Guan WJ, Ni ZY, Hu Y, Liang WH, Ou CQ, He JX, et al. Clinical Characteristics of Coronavirus Disease 2019 in China. N Engl J Med. 2020.

5. Yang X, Yu Y, Xu J, Shu H, Xia J, Liu H, et al. Clinical course and outcomes of critically ill patients with SARS-CoV-2 pneumonia in Wuhan, China: a single-centered, retrospective, observational study. Lancet Respir Med. 2020;8(5):475–81.

6. Zhang C, Wu Z, Li JW, Zhao H, Wang GQ. Cytokine release syndrome in severe COVID-19: interleukin-6 receptor antagonist tocilizumab may be the key to reduce mortality. Int J Antimicrob Agents. 2020;55(5):105954.

7. Nahum J, Morichau-Beauchant T, Daviaud F, Echegut P, Fichet J, Maillet JM, et al. Venous Thrombosis Among Critically Ill Patients With Coronavirus Disease 2019 (COVID-19). JAMA Netw Open. 2020;3(5):e2010478.

8. Ackermann M, Verleden SE, Kuehnel M, Haverich A, Welte T, Laenger F, et al. Pulmonary Vascular Endothelialitis, Thrombosis, and Angiogenesis in Covid-19. N Engl J Med. 2020.

9. Wynants L, Van Calster B, Bonten MMJ, Collins GS, Debray TPA, De Vos M, et al. Prediction models for diagnosis and prognosis of covid-19 infection: systematic review and critical appraisal. BMJ. 2020;369:m1328.

10. Zhou F, Yu T, Du R, Fan G, Liu Y, Liu Z, et al. Clinical course and risk factors for mortality of adult inpatients with COVID-19 in Wuhan, China: a retrospective cohort study. Lancet. 2020;395(10229):1054–62.

11. Yu T, Cai S, Zheng Z, Cai X, Liu Y, Yin S, et al. Association Between Clinical Manifestations and Prognosis in Patients with COVID-19. Clin Ther. 2020.

12. Wollenstein-Betech S, Cassandras CG, Paschalidis IC. Personalized Predictive Models for Symptomatic COVID-19 Patients Using Basic Preconditions: Hospitalizations, Mortality, and the Need for an ICU or Ventilator. medRxiv. 2020.

13. Fine MJ, Auble TE, Yealy DM, Hanusa BH, Weissfeld LA, Singer DE, et al. A prediction rule to identify low-risk patients with community-acquired pneumonia. N Engl J Med. 1997;336(4):243–50.

14. Government of Ontario. How Ontario is responding to COVID-19. Available via the Internet at https://www.ontario.ca/page/how-ontario-is-responding-covid-19#section-0. Last accessed June 10, 2020. Toronto, Canada: Queen’s Printer for Ontario; 2020.

15. Statistics Canada. Population estimates, quarterly. Table: 17-10-0009-01 (formerly CANSIM 051-0005). Available via the Internet at https://www150.statcan.gc.ca/t1/tbl1/en/tv.action?pid=1710000901. Last accessed May 29, 2020. 2020.

16. Neilsen K. A timeline of the novel coronavirus in Ontario. Available via the Internet at https://globalnews.ca/news/6859636/ontario-coronavirus-timeline/. Last accessed May 29, 2020. Global News. Toronto, Canada; 2020.

17. Tuite AR, Greer AL, De Keninck S, Fisman DN. Risk for COVID-19 Resurgence Related to Duration and Effectiveness of Physical Distancing in Ontario, Canada. Ann Intern Med. 2020.

18. Deeks SL, Lim GH, Walton R, Fediurek J, Lam F, Walker C, et al. Prolonged Pertussis Outbreak in Ontario Originating in an Under-immunized Religious Community. Can Commun Dis Rep. 2014;40(3):42–9.

19. Case Definition—Novel Coronavirus (COVID-19). Available via the Internet at http://www.health.gov.on.ca/en/pro/programs/publichealth/coronavirus/docs/2019_case_definition.pdf. Toronto, Canada: Public Health Ontario; 2020.

20. Fisman D, Lapointe-Shaw L, Bogoch I, McCready J, Tuite AR. Failing our Most Vulnerable: COVID-19 and Long-Term Care Facilities in Ontario. medRxiv 2020.04.14.20065557; doi: https://doi.org/10.1101/2020.04.14.20065557. 2020.

21. Hosmer DW, Lemeshow S. Applied Logistic Regression: John Wiley & Sons; 1989.

22. The Novel Coronavirus Pneumonia Emergency Response Epidemiology Team. Vital Surveillances: The Epidemiological Characteristics of an Outbreak of 2019 Novel Coronavirus Diseases (COVID-19) — China, 2020. Available via the Internet at http://weekly.chinacdc.cn/en/article/doi/10.46234/ccdcw2020.032. Last accessed April 30, 2020. China Centers for Disease Control Weekly. 2020;2(8):113–22.

23. Hou W, Zhang W, Jin R, Liang L, Xu B, Hu Z. Risk factors for disease progression in hospitalized patients with COVID-19: a retrospective cohort study. Infect Dis (Lond). 2020;52(7):498–505.

24. Caramelo F, Ferreira N, O’liveiros B. Estimation of risk factors for COVID-19 mortality - preliminary results. medRxiv 2020.02.24.20027268; doi:https://doi.org/10.1101/2020.02.24.20027268. Last accessed June 20, 2020.

25. de Lusignan S, Dorward J, Correa A, Jones N, Akinyemi O, Amirthalingam G, et al. Risk factors for SARS-CoV-2 among patients in the Oxford Royal College of General Practitioners Research and Surveillance Centre primary care network: a cross-sectional study. Lancet Infect Dis. 2020.

26. Gonzalez-Rubio J, Navarro-Lopez C, Lopez-Najera E, Lopez-Najera A, Jiminez-Diaz L, Navarro-Lopez JD, et al. Cytokine Release Syndrome (CRS) and Nicotine in COVID-19 Patients: Trying to Calm the Storm. Available via the Internet at https://doi.org/10.3389/fimmu.2020.01359. Last accessed June 14, 2020. Front. Immunol. 2020.

27. Hamer M, Kivimaki M, Gale CR, Batty GD. Lifestyle risk factors, inflammatory mechanisms, and COVID-19 hospitalization: A community-based cohort study of 387,109 adults in UK. Brain Behav Immun. 2020.

28. Leung JM, Sin DD. Smoking, ACE-2, and COVID-19: Ongoing Controversies. Eur Respir J. 2020.

29. Bai X, Fang C, Zhou Y, Bai S, Liu Z, Chen Q, et al. Predicting COVID-19 malignant progression with AI techniques. medRxiv 2020.03.20.20037325; doi: https://doi.org/10.1101/2020.03.20.20037325. Last accessed June 20,2020. 2020.

